# Adjustment for energy intake in nutritional research: a causal inference perspective

**DOI:** 10.1101/2021.01.20.21250156

**Authors:** Georgia D Tomova, Kellyn F Arnold, Mark S Gilthorpe, Peter WG Tennant

## Abstract

**Background:** Four models are commonly used to adjust for energy intake when estimating the causal effect of a dietary component on an outcome; (1) the ‘standard model’ adjusts for total energy intake, (2) the ‘energy partition model’ adjusts for remaining energy intake, (3) the ‘nutrient density model’ rescales the exposure as a proportion of total energy, and (4) the ‘residual model’ indirectly adjusts for total energy by using a residual. It remains underappreciated that each approach evaluates a different estimand and only partially accounts for proxy confounding by common dietary causes.

**Objective:** To clarify the implied causal estimand and interpretation of each model and evaluate their performance in reducing dietary confounding.

**Design:** Semi-parametric directed acyclic graphs and Monte Carlo simulations were used to identify the estimands and interpretations implied by each model and explore their performance in the absence or presence of dietary confounding.

**Results:** The ‘standard model’ and the mathematically identical ‘residual model’ estimate the *average relative causal effect* (i.e., a ‘substitution’ effect) but provide biased estimates even in the absence of confounding. The ‘energy partition model’ estimates the *total causal effect* but only provides unbiased estimates in the absence of confounding or when all other nutrients have equal effects on the outcome. The ‘nutrient density model’ has an obscure interpretation but attempts to estimate the average relative causal effect rescaled as a proportion of total energy intake. Accurate estimates of both the total and average relative causal effects may instead be estimated by simultaneously adjusting for all dietary components, an approach we term the ‘all-components model’.

**Conclusion:** Lack of awareness of the estimand differences and accuracy of the four modelling approaches may explain some of the apparent heterogeneity among existing nutritional studies and raise serious questions regarding the validity of meta-analyses where different estimands have been inappropriately pooled.

## Introduction

Estimating the causal effect of an individual dietary component on one or more health outcomes is a common practice in nutrition research. The purported aim is to identify foods or nutrients that are particularly beneficial or harmful to health, and hence reveal potential targets for public health or policy intervention. For example, many countries have introduced taxes on sugar-sweetened beverages because of their high concentration of non-milk extrinsic sugars and their estimated contribution to the risks of obesity and other adverse health outcomes.(1,2)

Randomised controlled trials are generally difficult to perform in larger samples over longer time periods, and the effects observed may not generalize to dietary practices in the target population.(3) Nutrition research is therefore highly reliant on the analysis of observational data, which brings several challenges for causal inference. One of the biggest of these is how to separate the effects of individual dietary components from the effects of the overall diet. Typically, those who consume a greater quantity of any one dietary component will also consume a greater overall quantity of food and have a greater overall energy intake.(4) Those who consume a greater overall quantity of food are often also systematically different in several other important ways, such as body size and composition.(4) Separating the effect of a single nutrient exposure from the effects of body size, body composition, metabolic efficiency, and overall energy intake, is however extremely challenging.(5) Identifying the exact physiological, psychological and sociocultural determinants of dietary intake and composition is not straightforward. Many of these determinants cannot be measured directly or are simply unknown. Overall energy intake is often the best available ‘proxy’ for these determinants, and is hence routinely used to address confounding.(4)

Several strategies have been proposed to ‘control’ (i.e. statistically adjust) for differences in overall energy intake when attempting to estimate the effects of individual dietary components, and there has been considerable debate about which of these strategies is most appropriate.(4–9) The two most common approaches are: the ‘**standard model**’, which involves adjusting for *total energy intake* (i.e. total intake of calories from all sources *including* the nutrient exposure of interest); and the ‘**energy partition model**’, which involves adjusting for the *remaining energy intake* (i.e. the intake of calories from all sources *excluding* the exposure nutrient of interest). A third approach, known as the ‘**nutrient density model**’, involves examining the nutrient exposure as a proportion (percentage) of total energy intake, with or without further adjustment for total energy intake. Finally, the ‘**residual model**’ involves adjusting for the residual produced by regressing the nutrient exposure on total energy intake.

Although ostensibly similar in purpose, the choice of energy adjustment strategies has important implications, for both the causal effect being targeted (i.e. the estimand) and the accuracy of the estimate obtained.(8) In theory, applied researchers are encouraged to select the adjustment strategy that is most compatible with their research question, or otherwise to carefully present and interpret different estimates.(8) However, in practice, there is often limited explicit justification given for the approach(es) adopted and reported. Even when the models are correctly interpreted, there is little or no explanation regarding which approach is most suitable, and instead several approaches are commonly used and compared, even when they target different causal effect estimands.(10,11)

In this study, we use directed acyclic graphs (DAGs) and simulations to clarify the estimand and appropriate causal interpretation for each adjustment strategy. We also explore the performance of each strategy for reducing confounding by common causes of dietary intake and composition. Throughout, we consider the illustrative example of the effect of non-milk extrinsic sugars (referred to as ‘sugar(s)’ for simplicity) on body weight.

## CONSIDERING COMPOSITIONAL NUTRITION DATA USING DAGS

A key challenge in the analysis of nutritional data is recognizing that total energy intake, together with energy intake from individual dietary components, represents an example of compositional data.(12) Data are compositional when a ‘whole’ variable (i.e. total energy intake) can be divided into meaningful ‘part’ variables which together sum to that whole (i.e. energy intake from individual dietary components).(13) Although we might consider these whole and part variables to be distinct, in reality they represent the same variable at different levels of aggregation.

Compositional data can be depicted using a type of semi-parametric DAG, as introduced by Arnold et al.(14). DAGs are causal diagrams in which variables (nodes) are connected by unidirectional arrows (arcs) to depict hypothesized causal relationships between them; no node may indirectly cause itself.(15) Although they are generally used to depict probabilistic relationships, they can also be used to depict situations for which the value of one variable is completely determined by one or more parent variables(16). We depict probabilistic and deterministic variables with single and double-outlined rectangles, respectively, and probabilistic and deterministic relationships with single and double-lined arrows, respectively. We also place a dashed box around the compositional variables to highlight that they occur at the same point in time.(14) The DAG in **Figure 1** depicts our illustrative scenario, where the energy intake from sugars and all other energy sources completely determines total energy intake and (probabilistically) affects body weight.

**Figure 1.**
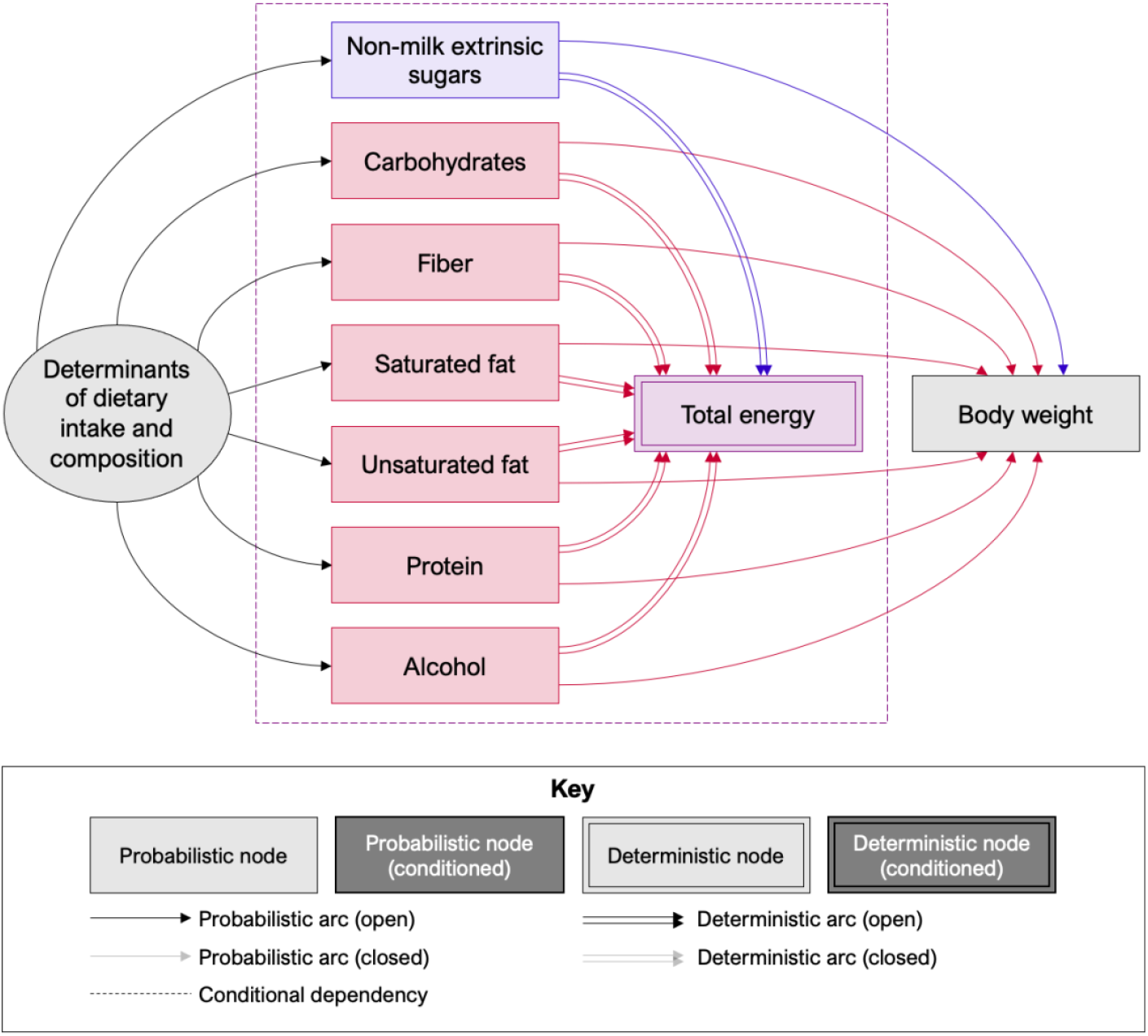
Directed acyclic graph illustrating the compositional nature of nutritional data. Total energy intake (the ‘whole’, purple) is fully determined by energy intake from seven constituent macronutrients (the ‘parts’, blue and red). The nutrient exposure (non-milk extrinsic sugars, blue) and six competing energy sources (red) all cause the outcome (body weight, grey) and are themselves caused by all unobserved determinants of dietary intake and composition (grey).

Depicting the relationship in this way – with total energy intake as the ‘consequence’ of the energy intake from all individual sources – is helpful for understanding the impact of different adjustment strategies because total energy intake can be conceptualized as a ‘collider’. A collider is a variable that is simultaneously caused by two or more other variables.

Where the collider is *completely determined* by its individual components, as in the case of total energy intake, conditioning on the collider (i.e., holding its value fixed) creates a dependency between all the constituent components, such that any change in the value of one component must be accompanied by an equal but opposite average change in all other (unconditioned) components. Analyses that adjust for total energy intake therefore evaluate the ‘substitution’ effect of exchanging the exposure with one or more other component nutrients.(14) The compositional nature of the data should be recognized when selecting the most appropriate adjustment strategy.

## TOTAL VERSUS RELATIVE CAUSAL EFFECT ESTIMANDS

Several authors have examined the interpretation of the different approaches to energy adjustment,(4,6–8) but none have explicitly considered the target estimand of each approach. This is likely because none of the models were developed within a formal causal framework. The estimand and estimating performance of each approach must therefore be inferred from theory.

The **total causal effect** of a nutrient exposure (e.g. sugars) on a health outcome (e.g. body weight), is the individual effect of increasing energy intake from that exposure while keeping all other sources of energy intake constant; this has previously been described as an ‘additive’ effect.(8) Since total energy intake is a ‘collider’ between the exposure nutrient and the outcome, it should be evident that an unbiased estimate of this effect cannot be obtained by adjusting for total energy intake. Therefore, of the four most common adjustment approaches, only the energy partition model targets this effect. This model can be expected to improve the precision of the estimate (by reducing unexplained heterogeneity in the outcome) and reduce confounding bias from common causes of diet (**Figure 2a**). However, this strategy would not be expected to fully eliminate confounding by common causes of diet if the competing dietary components have distinct effects on the outcome, since only their average effect will be adjusted for. In this instance, to target the total causal effect, adjusting simultaneously for each remaining dietary component – an approach we term the ‘**all-components model**’ – can be expected to provide a less biased estimate of this effect than would be obtained by adjusting for the average remaining energy intake (**Figure 2b**).

**Figure 2.**
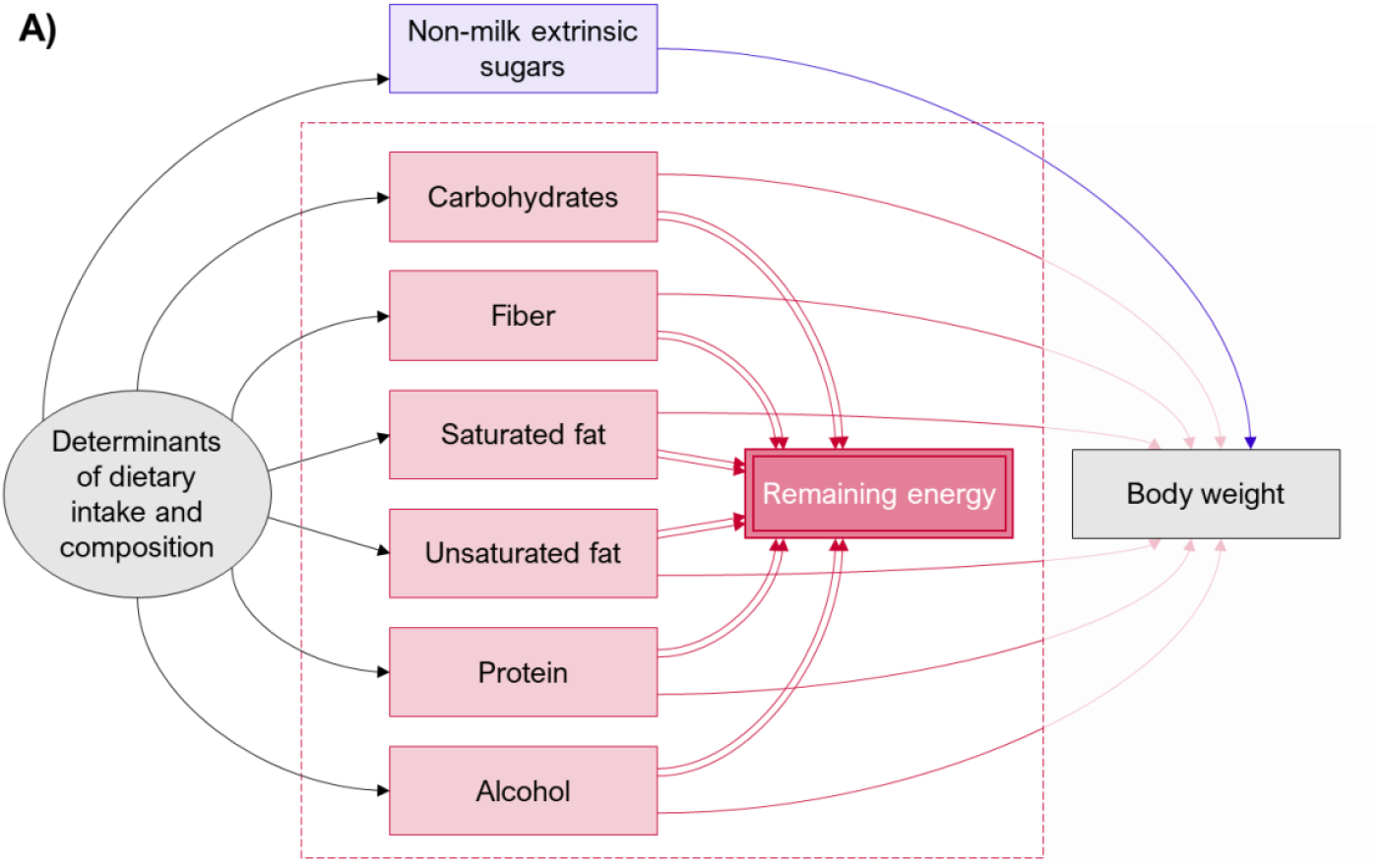
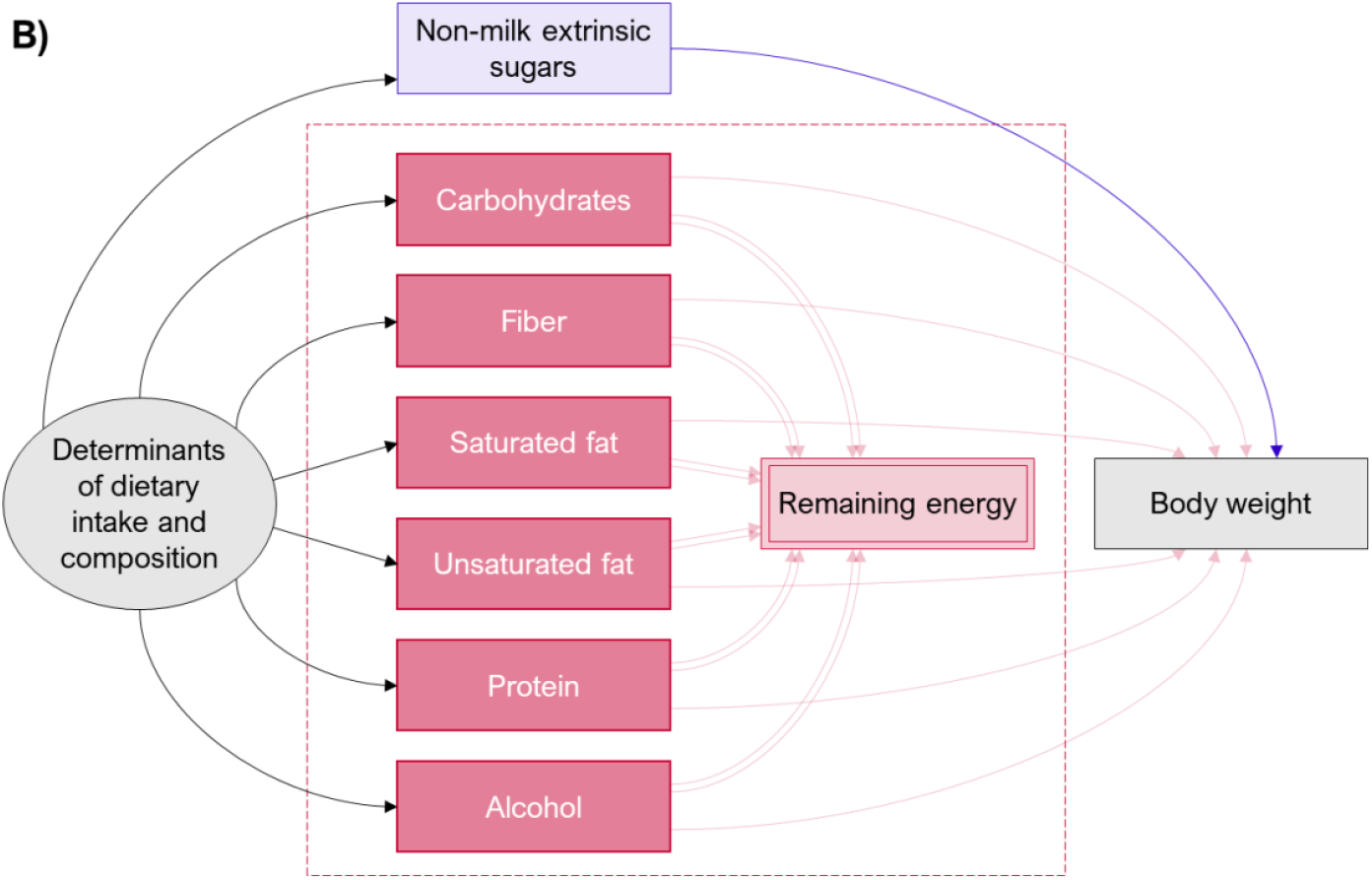
Directed acyclic graphs illustrating how confounding by common determinants of dietary intake and composition can be reduced when estimating the total causal effect (blue arc) of a nutritional exposure (e.g., non-milk extrinsic sugars, blue) on an outcome (e.g., body weight). Confounding by common determinants of dietary intake and composition will exist if one or more of the competing nutritional components (red) also cause the outcome (red arcs). This can be reduced by conditioning on the remaining energy intake (as show in A) or by conditioning on each of the competing nutritional components directly (as shown in B). For key see Figure 1.

A **relative causal effect** of a nutrient exposure is the *joint* effect of increasing energy intake from that nutrient while decreasing energy intake from one or more other energy sources to keep the total energy intake constant. In theory, there are many different relative causal effects that might be considered; requiring many different adjustment strategies. The **average relative causal effect** (also known as the weighted average compositional effect)(17) is the effect of a nutrient exposure relative to the weighted average effect of all other sources of energy, and is commonly estimated by adjusting for total energy (**Figure 3**). Other more specific average relative causal effects can also be targeted by additionally adjusting for specific competing sources of energy intake to remove them from the substitution group. However, as with remaining energy intake, adjusting for total energy intake is susceptible to residual confounding by common causes of diet if each residual dietary component has a distinct effect on the outcome. In this instance, a less biased estimate of the average relative causal effect may again be derived from the ‘all-components model’ (Figure 2b), by subtracting the weighted average effect of all the residual sources of energy intake from the total causal effect of the exposure.

**Figure 3.**
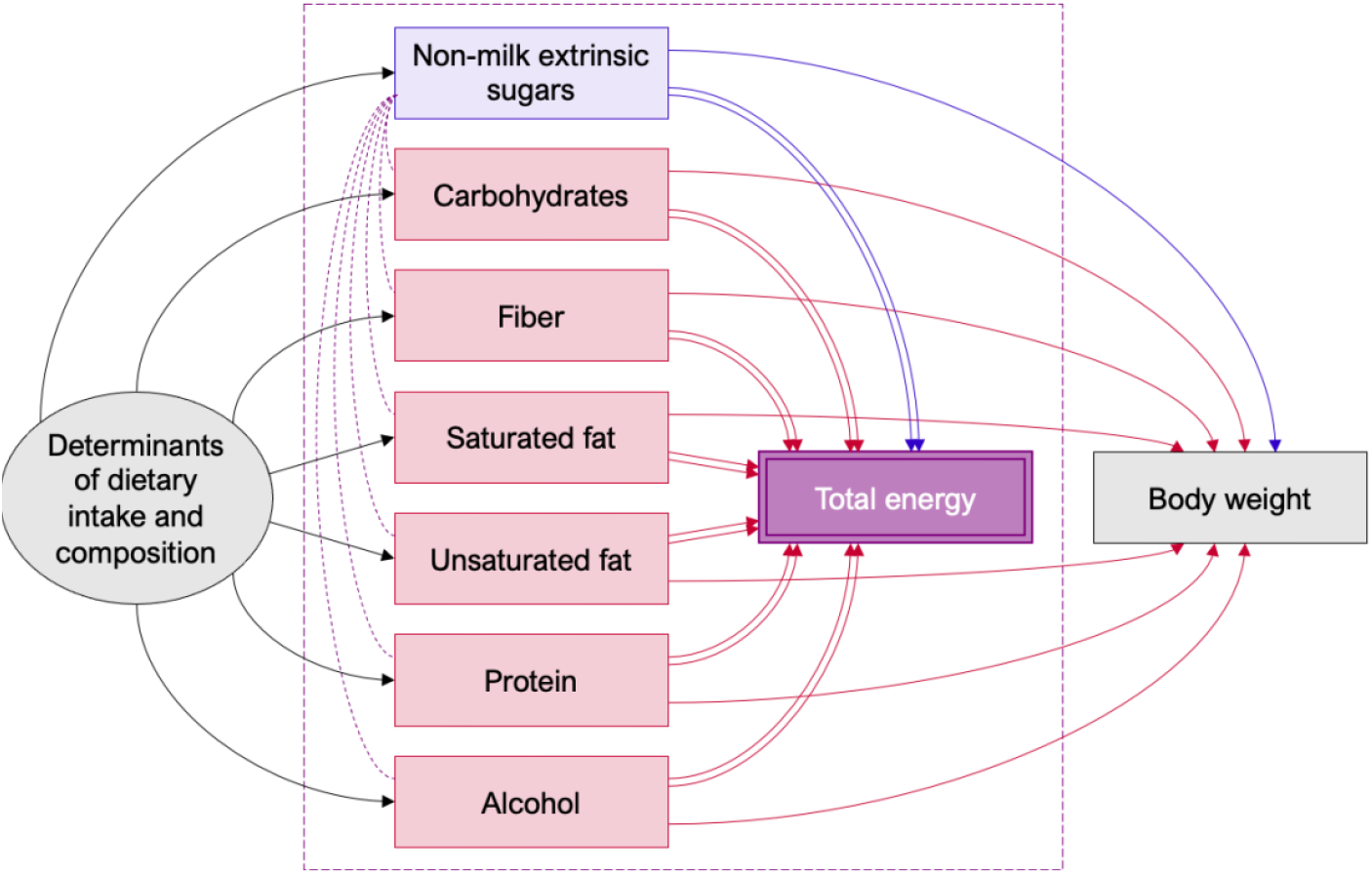
Directed acyclic graphs illustrating the consequences of adjusting for total energy intake when estimating the causal effect of a nutrient exposure (e.g., non-milk extrinsic sugars, blue) on an outcome (e.g., body weight, grey). Total energy is completely determined by the exposure nutrient (non-milk extrinsic sugars, blue) and all competing energy sources (red). Adjusting for total energy intake (purple) opens conditional dependencies between the exposure and all competing energy sources (purple dashed arcs), so that total causal effect (blue arc) is now in competition with the (average) effect of all competing energy sources (red arcs). The average relative causal effect thus represents the difference between the total causal effect of the exposure and the weighted average effect of all other energy sources. For key, see Figure 1.

## ILLUSTRATIVE EXAMPLE

To illustrate these principles, we explore the target estimand and the performance in *estimating* these quantities of the four standard energy adjustment strategies, the ‘all-components model’, and a reference unadjusted model, using simulated data. We consider the example of estimating the effect of sugar on body weight in a simple scenario where body weight is caused by the intake of seven macronutrients (including sugar), either in the presence or absence of confounding by common causes of dietary intake and composition. We only consider confounding that acts through the individual dietary components (i.e., no direct effect on the outcome) because this best illustrates the use of energy intake as a proxy of dietary determinants.

## Methods

Standardized data were simulated using the ‘dagitty’ (0.2-3) R package(18) to reflect the data generating process depicted in **Supplementary Figure 1**, where total energy intake was fully determined by the energy intake from seven macronutrients: 1) sugars, 2) carbohydrates, 3) fiber, 4) saturated fat, 5) unsaturated fat, 6) protein, and 7) alcohol. Total energy and remaining energy intake were not directly simulated. Instead, they were calculated from the sum of all macronutrient energy variables, or the sum of all energy variables except sugar, respectively. Each macronutrient was assigned a unique effect on body weight. Specific path coefficient values were chosen to represent plausible causal effects, and simulated variables were rescaled with plausible mean and standard deviation values informed by the National Diet and Nutrition Survey (see **Supplementary Table 1**).(19) All simulations and models (see below) were repeated in the presence of a single variable (U) that causes the intake of all macronutrients, to demonstrate the influence of confounding by common causes of dietary composition. Each simulation included 1,000 observations and was repeated over 100,000 iterations. We report the median effect estimate and 2.5th and 97.5th centiles (representing 95% simulation interval, SI) from the 100,000 iterations for each model. For ease of illustration, effect estimates are presented in kilograms per 100 kilocalories (kg/100kcal).

### Simulated Effects

The total causal effect is the effect of increasing energy intake from the exposure of interest (i.e., sugars) while maintaining the same levels of energy intake from all other sources. We simulated a total causal effect of 5kg/100kcal, meaning that body weight increased by an average of 5kg for each additional 100kcal of sugars consumed.

The average relative causal effect is the effect of increasing the energy intake from the exposure (i.e., sugars) while decreasing the energy intake from all other macronutrients to maintain the same total energy intake.

We simulated an average relative causal effect of 2kg/100kcal (equivalent to 0.4kg per 1% of total energy intake), meaning that body weight increased by an average of 2kg for each additional 100kcal (or 0.4kg for each additional 1% of total energy) derived from sugars rather than from other macronutrient sources.

### Models Examined

We present and discuss the results obtained from using the following six models.

#### 0 The unadjusted model

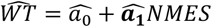

This model targets the *total causal effect* of sugars (*MNES*) on body size (*WT*). Because the model does not adjust for energy intake or any other variables, the coefficient 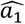 should produce an unbiased estimate of the desired estimand only where there exists no confounding by common causes of diet.

#### 1 The energy partition model

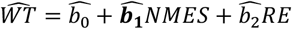

This model also targets the *total causal effect* of sugars (*MNES*) on body weight (*WT*), and attempts to minimize confounding by common causes of diet by adjusting for remaining energy intake (*RE*) (i.e., total intake of calories from all sources *excluding* sugars). Where there exists such confounding, the coefficient 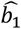 is therefore expected to produce a less-biased estimate of the desired estimand than in the unadjusted model (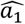 in model 0); however, residual confounding may remain where competing nutrients have different effects on the outcome.

#### 2 The standard model

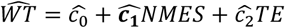

This model targets the *average relative causal effect* of sugars (*MNES*) on body weight (*WT*), and attempts to minimize confounding by common causes of diet by adjusting for total energy intake (*TE*) (i.e., total intake of calories from all sources *including* sugars). The coefficient 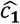 is expected to provide an unbiased estimate of this effect in the absence of confounding. The effect may be biased in the presence of confounding; residual confounding may also remain where competing nutrients have different effects on the outcome.

#### 3 The nutrient density model

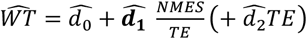

This model involves transforming the nutrient exposure into a proportion (percentage) of total energy intake; additional adjustment may also be made for total energy intake (*TE*), an approach that has been termed the ‘*multivariable* nutrient density model’.(4) The estimand targeted by this model is unclear, but it is plausibly an attempt to estimate the *average relative causal effect* of sugars (*MNES*) on body weight (*WT*), rescaled as a percentage of total energy intake (i.e. 0.4kg/1%). However, the coefficient 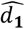 represents an obscure quantity that conflates both the effect of the nutrient exposure and of the reciprocal of total energy intake, and it is therefore expected to be biased regardless of confounding by common causes of diet. We present models with (*3a*) and without (*3b*) adjustment for total energy intake (*TE*).

#### 4 The residual model

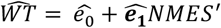

This two-stage approach involves regressing the nutrient exposure on total energy intake 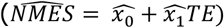, and entering the model residual 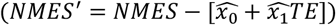 into a second unadjusted model. The approach is mathematically identical to the standard model, and therefore also targets the *average relative causal effect* of sugars (*MNES*) on body weight (*WT*). Like in the standard model, the coefficient 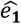 is expected to provide an unbiased estimate in the absence of confounding; however, the effect may be biased in the presence of confounding or residual confounding if the remaining nutrient components have distinct effects on the outcome.

#### 5 The all-components model

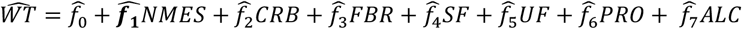

This model targets the *total causal effect* of sugars (*MNES*) on body weight (*WT*) by adjusting for all individual component sources of energy: carbohydrates (*CRB*); fiber (*FBR*); saturated fat (*SF*); unsaturated fat (*UF*); protein (*PR*); alcohol (*ALC*). The coefficient estimate 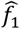 is expected to provide an unbiased estimate of this effect regardless of confounding by common causes of diet.

An unbiased estimate of the *average relative causal effect* (say, *ĝ*_1_) can also be estimated using this model. This is achieved by subtracting a weighted average of the estimated effects of all other individual component sources of energy from the total causal effect of the exposure (i.e., 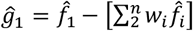, where *w*_*i*_ is the proportion of the remaining energy intake contributed by each component *i* = {2, …, *n*}).

## Results

Full results from the six models considered are given in **Table 1**.

**Table 1.**
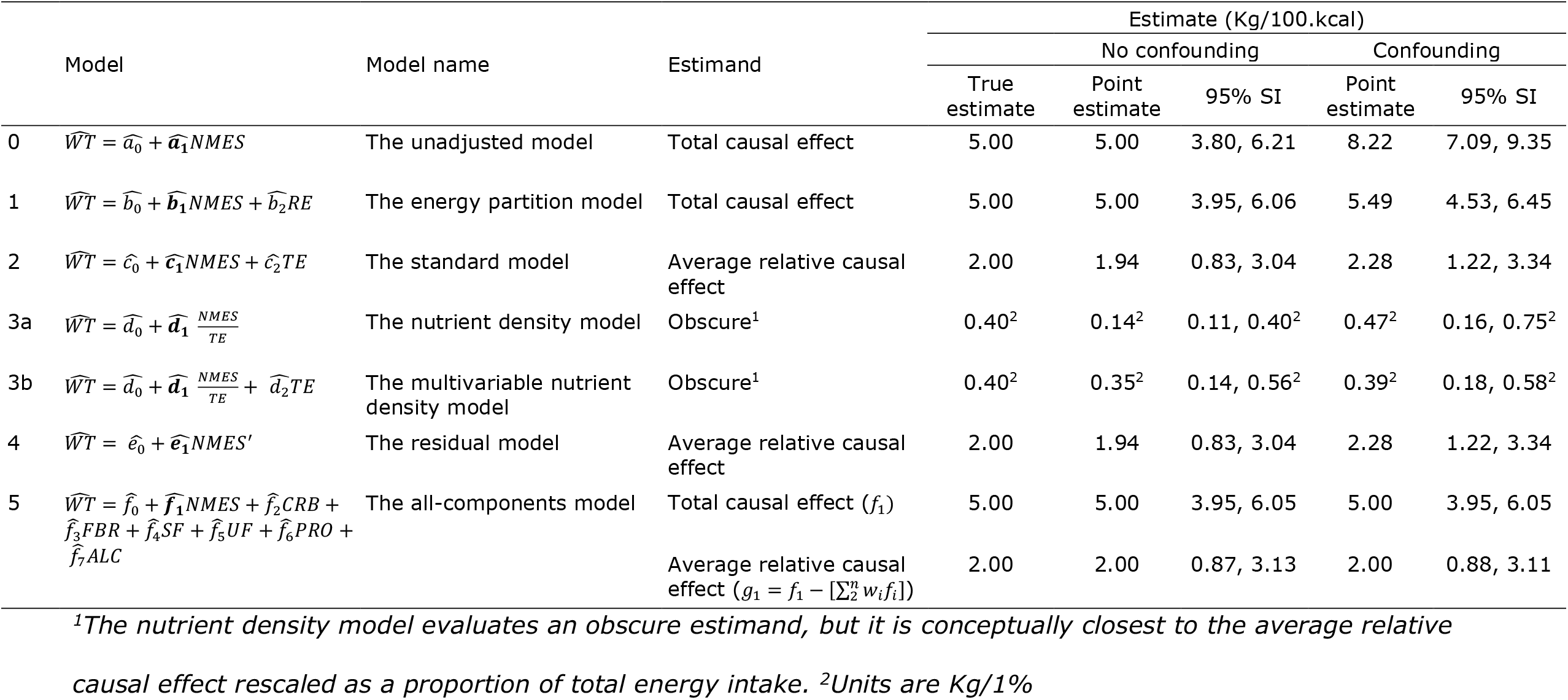
Regression coefficients (and 95% simulation intervals, SI) for the effect of sugars on body weight estimated using four different causal estimand scenarios and adjustment approaches.

### 0 The unadjusted model

With no confounding by common causes of diet, the unadjusted model returns an unbiased estimate of the total causal effect 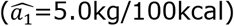. However, because the unadjusted model does not account for any competing sources of energy intake, the unadjusted model returns a severely biased estimate 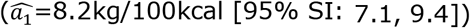 when there is confounding.

### 1 The energy partition model

With no confounding, the energy partition model returns an unbiased estimate of the total causal effect 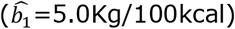. With confounding, the energy partition model returns a more accurate (but still biased) estimate (*b*_1_ = 5.5 [95 SI: 4.5, 6.5]), compared to the unadjusted model.

### 2 The standard model

With no confounding, the standard model returns a slightly biased estimate of the average relative causal effect 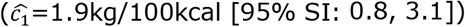. With confounding, the standard model also returns a biased estimate of the average relative causal effect 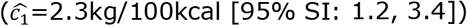.

### 3 The nutrient density model

a. With no confounding, the nutrient density model returns a severely biased estimate of the average relative causal effect 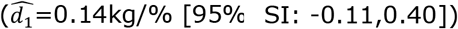. With confounding, the nutrient density model also returns a biased estimate of the average relative causal effect 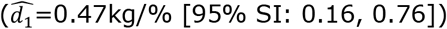.
b. With no confounding, the multivariable nutrient density model returns a more accurate estimate than the (unadjusted) nutrient density model, but one which is still biased 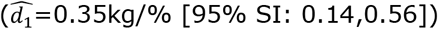. With confounding, the multivariable nutrient density model also returns a more accurate estimate than the (unadjusted) nutrient density model, but the estimate is still biased 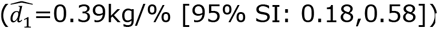.

### 4 The residual model

With no confounding, the residual model returns the same (biased) estimate of the average relative causal effect as the standard model 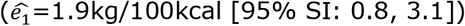. With confounding, the residual model also returns the same (biased) estimate of the average relative causal effect as the standard model 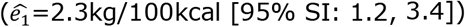.

### 5 The all-components model

With no confounding, the all-components model returns unbiased estimates of the total causal effect 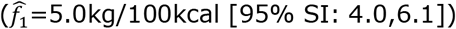 and average relative causal effect 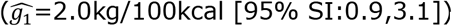. Because this model involves adjusting for all remaining component sources of energy separately, in the presence of confounding by common causes of diet it also returns unbiased estimates of the total causal effect 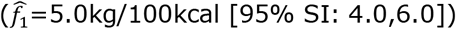 and average relative causal effect 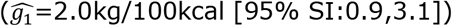.

## DISCUSSION

### Overview

This study used DAGs and simulations to explore the target estimands and performance of the four standard approaches to adjusting for energy intake in nutritional research, as well as a fifth model that involves adjusting for all individual energy components. We demonstrate that the four standard approaches evaluate different estimands with different interpretations, and that none of the four methods provide robust estimates in the presence of confounding by common causes of dietary intake and composition. In contrast, the ‘all-components’ model offers an accurate means to estimating both the total causal effect and average relative causal effect. The nutrient density model conflates the effects of the exposure and total energy and does not target a meaningful estimand, regardless of bias.

### Principal findings

Our simulations highlight four important considerations for the analysis and interpretation of nutritional data.

First, different modelling strategies target different estimands. The energy partition model is the only one of the four standard approaches that targets the total causal effect of the nutrient exposure on the outcome (i.e., the ‘additive’ effect of the exposure on top of the existing diet). In contrast, the standard model and the mathematically equivalent residual model both target the average relative causal effect of the nutrient exposure (i.e., the effect of ‘substituting’ the exposure with other calorific sources to maintain the same total energy intake). The nutrient density model targets an obscure estimand that conflates the effects of the nutrient exposure and the (inverse) effect of total energy intake; this is conceptually closest to the average relative causal effect, rescaled as a proportion of the total energy intake.

Second, we show that none of the four standard approaches provide robust estimates of their respective causal estimands in the presence of confounding by common causes of dietary intake and composition. There are different reasons for this which depend on the model used. For example, *the standard model* and the *energy partition model* only remove the average effect of energy intake. Residual confounding will therefore remain wherever the effect varies between different component energy sources. This assumption is, however, fundamental to the conduct of nutritional research, as there would otherwise be no justification for estimating the causal effect of one or more individual nutrient exposures. *The residual model* is algebraically identical to the standard model,(6) and as such suffers the same problems while offering no additional benefits.

*The nutrient density model* involves evaluating a proportion of energy intake (i.e., a ratio) as the exposure, rather than the absolute amount of the nutrient exposure consumed. For a variable expressed as a ratio, the individual causal effects of the constituent components cannot be separated and interpreted on their own. Ratio variables like these have obscure interpretations and are not robust to confounding bias.(20) Adjusting for total energy intake - as in the *multivariable nutrient density model* - offers considerably more accurate estimates of average relative causal effects by reducing confounding and reducing the distorting joint effects of the total energy denominator, but nevertheless remains biased.

Third, even in the absence of confounding, both the standard model and residual model returned slightly biased estimates (most probably due to a loss of information when combining the exposure and all remaining components into a single ‘total’ variable), and the nutrient density model produced moderately to severely biased estimates depending on whether additional adjustment was made for total energy intake.

Finally, we show that a model that includes all individual dietary components (i.e., *the all-components model*) offers a robust means to estimating both the total causal effect and average relative causal effect.

### Implications

None of the most common approaches to adjusting for energy intake in nutritional research provide robust estimates of meaningful causal effects. For some models, this is true even with no confounding by common causes of diet, reflecting fundamental issues with these approaches. This has serious implications for the validity and interpretation of existing studies that have used these models.

It remains underappreciated that adjusting for total energy intake and adjusting for remaining energy intake evaluate very different causal estimands. In our simulation, the true total causal effect of sugars on body weight was 5kg/100kcal, and the true average relative causal effect was 2kg/100kcal. These two estimands relate to very different questions that require very different interpretations. If this distinction is not recognized, there is a high chance of misinterpretation and confusion.

Unfortunately, meta-analyses of dietary exposures rarely separate studies based on their target estimand and/or modelling strategy(1), resulting in confusing summary estimates that are difficult - if not impossible - to interpret causally. The inappropriate synthesis of estimates from different estimands may therefore render many meta-analyses as meaningless.

Residual confounding is also likely to contribute to the heterogeneity of estimates observed in the literature, given the inadequacy of adjusting for energy intake using any of the traditionally recommended approaches.

More robust estimates can be obtained by adjusting simultaneously for all dietary components (as in the ‘all-components model’), but this is not common practice. However, this strategy does introduce a trade-off between minimizing bias (by including the largest number of components at the finest level of detail) and maximizing precision (by having to estimate many parameters, i.e., one for each additional dietary component). Creating and adjusting for latent dietary profiles may offer a more parsimonious and hence more efficient approach to adjusting for common dietary causes, but there is still likely to be some accuracy trade-off.(21)

### Recommendations

Studies that seek to estimate the causal effect of one or more dietary components on one or more outcomes should clearly state their target estimand(s) of interest and justify an adjustment strategy for estimating this effect.

Meta-analyses should only attempt to pool estimates for identical estimands, and extra care should be taken by reviewers to determine the implied estimand where this is not explicit. DAGs offer a simple means to identifying the appropriate adjustment set for a particular estimand, and guidelines are now available on how best to report their use.(22)

A single model that includes *all* individual components of the diet may provide the simplest and most accurate approach to estimating both the total causal effect and any relative causal effect of interest.

### Strengths and weaknesses

While useful for demonstrating theoretical concepts, data simulations are over-simplifications of reality. The true causal effect of sugar intake, and of all other macronutrients, on weight are likely to differ from what was simulated. The specific values were selected to most clearly illustrate the issues at hand, and no effects reported in this study should be interpreted in the nutrition domain. Weight and all dietary variables were simulated to be multivariate normal, which does not reflect reality. To aid demonstration, we transformed these variables to have plausible means and standard deviations based on observations in official data sources, though this has no substantive impact on the results derived. Although we reduced the standard deviations to minimize the occurrence of negative values, some negative and biologically implausible values were nevertheless simulated in some instances which, although nonsensical, did not impact the validity of the simulations or the interpretation of results.

When the aim is to reduce confounding, overall energy intake is adjusted as a proxy of unobserved determinants of dietary intake and composition. Therefore, in this study we only considered proxy confounding by common causes of diet. Minimizing confounding by common causes of diet does not eliminate the need for a carefully considered adjustment set. In the presence of standard confounding (i.e., variables that cause both the exposure and the outcome), adjusting for all dietary components would not be sufficient to eliminate all confounding bias. In such circumstances, when estimating the causal effect of a nutrient exposure on a health outcome, the complete adjustment set should be carefully selected and justified, ideally using DAGs.(22)

## CONCLUSION

It is not fully appreciated that the most common approaches to adjusting for energy intake in nutritional research target different estimands with different interpretations. Moreover, none of these approaches offer complete adjustment for confounding from common causes of dietary intake and composition. These two issues together may explain a large portion of the heterogeneity in effect estimates between nutritional studies. The alternative single model that includes *all* individual components of the diet may provide the simplest and most accurate approach to estimating total causal effects and any desired relative causal effects.

## Data Availability

Data described in the manuscript, code book, and analytic code will be made publicly and freely available without restriction at: https://github.com/georgiatomova/adjustment-energy-intake

## AUTHOR CONTRIBUTIONS

GDT, MSG, and PWGT conceived and designed the study. GDT conducted the data simulations. All authors were involved in the interpretation of the simulation results. GDT drafted the manuscript and all authors contributed to critically reviewing and revising the draft. All authors read and approved the final manuscript before submission and agreed with the decision to submit to *medRxiv*.

## SUPPLEMENTARY MATERIALS

**Supplementary Table 1.** Target mean and standard deviation values of the variables in the simulated data.

| Variable (unit) | Mean (SD) |
| --- | --- |
| Weight (kg) | 80 (25) |
| Sugars (kcal) | 250 (125) |
| Carbohydrates (kcal) | 500 (250) |
| Fiber (kcal) | 100 (50) |
| Saturated fat (kcal) | 275 (125) |
| Unsaturated fat (kcal) | 400 (200) |
| Protein (kcal) | 300 (150) |
| Alcohol (kcal) | 175 (50) |
| Total energy intake (kcal) | 2000 (400) |
| Remaining energy intake (kcal) | 1750 (400) |

**Supplementary Figure 1.**
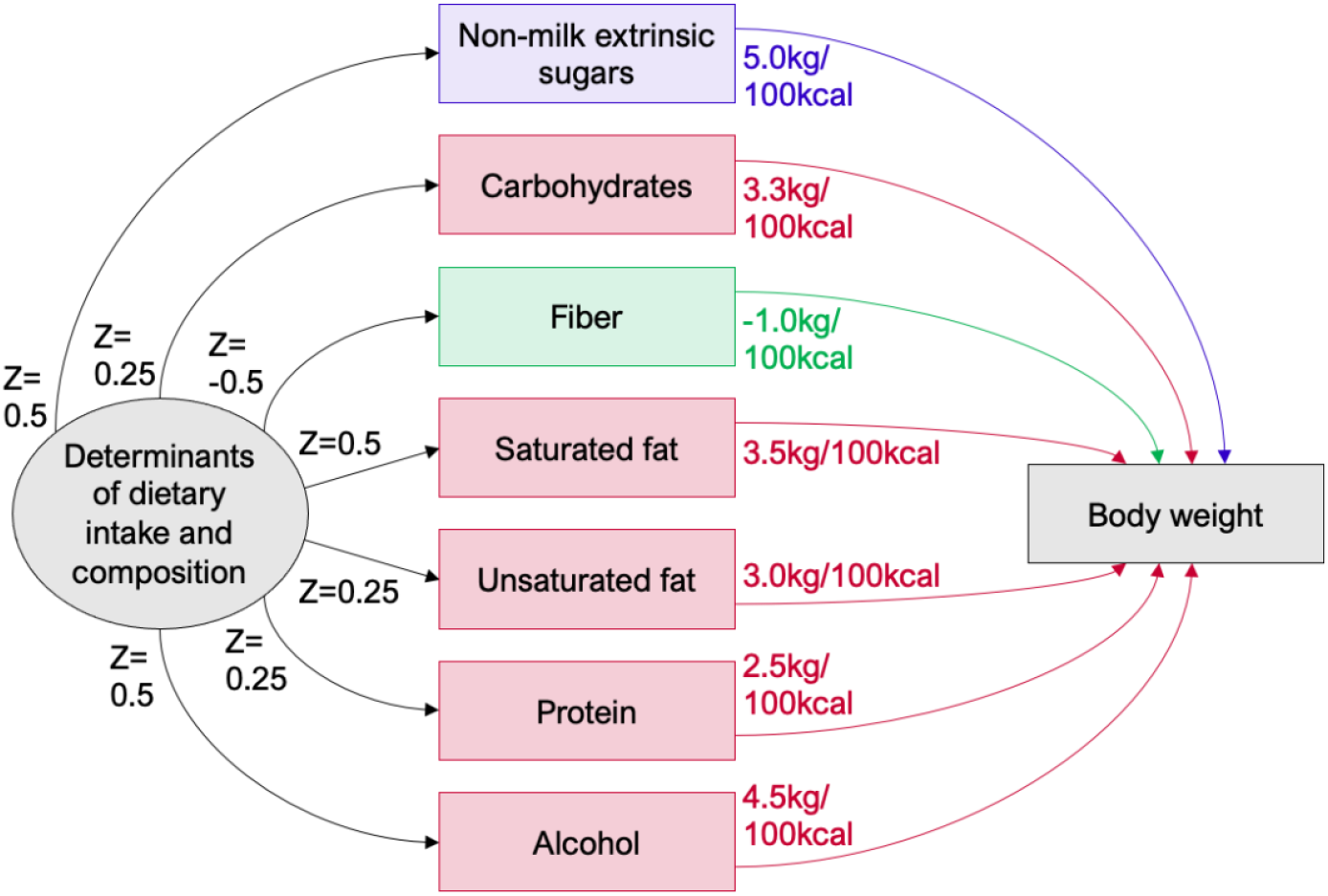
Path diagram of the simulated data structure and path coefficients. Energy from sugars was assigned a standardized path coefficient of 0.25, carbohydrates 0.33, fiber −0.02 (because the calorific energy of insoluble fiber is not obtainable), saturated fat 0.175, unsaturated fat 0.24, protein 0.15, and alcohol 0.09.

